# Detection of acne by deep learning object detection

**DOI:** 10.1101/2021.12.05.21267310

**Authors:** Amandip Sangha, Mohammad Rizvi

**Affiliations:** Askin; Oslo University Hospital Rikshospitalet

## Abstract

**Importance:** State-of-the art performance is achieved with a deep learning object detection model for acne detection. There is little current research on object detection in dermatology and acne in particular. As such, this work is early in this field and achieves state of the art performance.

**Objective:** Train an object detection model on a publicly available data set of acne photos.

**Design, Setting, and Participants:** A deep learning model is trained with cross validation on a data set of facial acne photos.

**Main Outcomes and Measures:** Object detection models for detecting acne for single-class (acne) and multi-class (four severity levels). We train and evaluate the models using standard metrics such as mean average precision (mAP). Then we manually evaluate the model predictions on the test set, and calculate accuracy in terms of precision, recall, F1, true and false positive and negative detections.

**Results:** We achieve state-of-the art mean average precision mAP@0.5 value of 37.97 for the single class acne detection task, and 26.50 for the 4-class acne detection task. Moreover, our manual evaluation shows that the single class detection model performs well on the validation set, achieving true positive 93.59 %, precision 96.45 % and recall 94.73 %.

**Conclusions and Relevance:** We are able to train a high-accuracy acne detection model using only a small publicly available data set of facial acne. Transfer learning on the pre-trained deep learning model yields good accuracy and high degree of transferability to patient submitted photographs. We also note that the training of standard architecture object detection models has given significantly better accuracy than more intricate and bespoke neural network architectures in the existing research literature.

**Key Points:** *Question:* Can deep learning-based acne detection models trained on a small data set of publicly available photos of patients with acne achieve high prediction accuracy?

*Findings:* We find that it is possible to train a reasonably good object detection model on a small, annotated data set of acne photos using standard deep learning architectures.

*Meaning:* Deep learning-based object detection models for acne detection can be a useful decision support tools for dermatologists treating acne patients in a digital clinical practice. It can prove a particularly useful tool for monitoring the time evolution of the acne disease state over prolonged time during follow-ups, as the model predictions give a quantifiable and comparable output for photographs over time. This is particularly helpful in teledermatological consultations, as a prediction model can be integrated in the patient-doctor remote communication.

## Introduction

Acne is a chronic inflammatory disease of the skin and is among the most common skin conditions^5^ in the world. The Global Burden of Disease study (2010) found that acne was the eight most prevalent disease globally, estimated to affect 9.4% of the global population^1^. More than 95% of teenage boys and more than 85% of teenage girls are affected, and more than 20% of those affected have moderate to severe acne, and 50% continue to suffer from acne in their adulthood.

The pathogenesis of acne is multifaceted. Hyperkeratinization is the excessive accumulation of dead skin cells causing a blockage of hair follicles. Abnormal keratinization of the infundibular epitheleum causes an obstruction of sebaceous follicles. Androgenic stimulation of sebaceous glands increase secretion. Microbial colonization of pilosebaceous units by the bacteria *cutibacterium acnes* contributes to inflammation. The clinical and thus visual presentation of acne results in

▪ comedones - whiteheads and blackheads (mild)
▪ pustules
▪ papules
▪ nodules and cysts (severe)

We train a deep learning object detection model on a publicly available image set to predict acne in photos. Our model achieves state of the art accuracy with an existing deep neural network architecture and no special preprocessing or hyperparameter evolution. We train on both single class (acne) detection and multi class (severity levels 1-4).

## Methods

### Data

We train the model on publicly available data set ACNE04^3^ by Wu et al (ref). This data set consists of 1457 photographs of patients. The photos are taken of the patients’ faces at an approximate angle of 70 degrees from the front. The photos have been manually annotated by experts, so that the acne lesions in the photographs have been enclosed in bounding boxes, a total number of 18983 bounding boxes.

We split the data set randomly into a training-test split of 80% for training (1165 photos, 14879 lesion bounding boxes) and the remaining 20% for testing (292 photos, 4104 lesion bounding boxes).

The ACNE04 dataset is used for training and validating the model, i.e. building the deep learning model and deriving model validation metrics such as precision, recall and mAP. Secondarily, we employ a small data set consisting of photographs submitted by patients in our clinical practice. We use only completely anonymous photographs where the person is unidentifiable.

### Deep learning object detection model

Object detection is the branch of computer vision that deals with the identification of specific objects within digital photos. Deep learning-based models have achieved great success in the field of object detection recently and is now the dominant technique. These models are deep convolutional neural networks^17^ which are trained on large, annotated image datasets to predict rectangular bounding boxes around objects of interest in the images.

We use the model *YOLOv5* which is a convolutional neural network consisting of 773 layers and 141.8 million parameters. Recall that the YOLO-family of object detection models is an example of a *single-stage detector* where a single neural network is responsible for predicting bounding boxes and class probabilities from full images directly in one evaluation. Object detection is framed as a regression problem of spatially separated bounding rectangles. This is in contrast to other prominent object detection models that are *two-stage detectors*, for example the Fast R-CNN family of models which employ one neural network for proposing regions of interest and another neural network for predicting bounding boxes within the proposed regions.

The YOLOv5 P6^15^ model for object detection is a single-stage detector that has been pre-trained on the COCO^9^ data set.

| Model | YOLOv5x6 |
| --- | --- |
| Image size (px) | 1280 |
| mAP<sup>val</sup><br>0.5:0.95 | 54.4 |
| mAP<sup>test</sup><br>0.5:0.95 | 54.4 |
| mAP<sup>val</sup><br>0.5 | 72.0 |
| Speed<br>V100 (ms) | 22.4 |
| params<br>(M) | 141.8 |
| FLOPS<br>(B) | 222.9 |

We use default hyperparameter values: learning rate 0.01, SGD momentum 0.937 and weight decay 0.0005, and conduct no hyperparameter evolution.

Recall the standard evaluation metrics of the object detection task. Intersection over Union, *IoU*, is the degree of overlap of two enclosed pixel regions. This clearly measures the accuracy of a predicted rectangle.

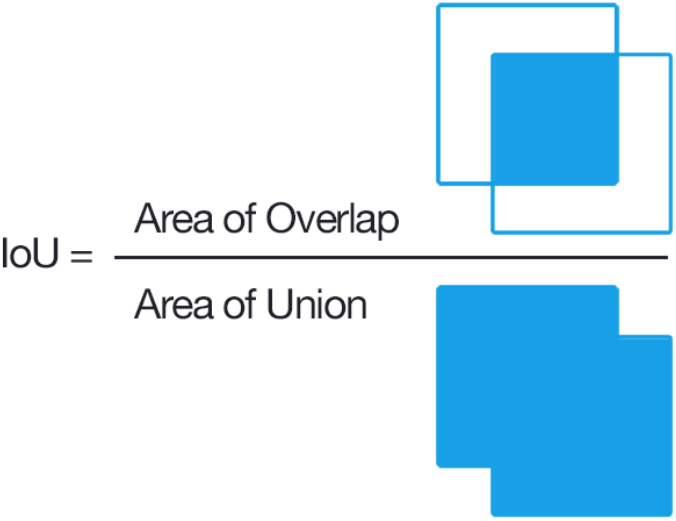

*Precision* and *recall* should be familiar notions from binary classification. *Precision* is the fraction of correct detections among all detections

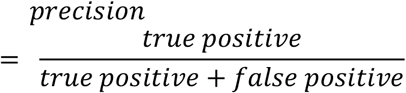

*Recall* is the fraction of correct detections retrieved

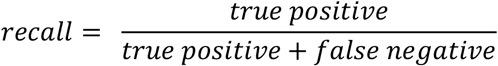

By plotting the precision and recall as a curve in the plane, one gets the Precision-Recall-curve. The area under this curve is referred to as *average precision* (AP). The *mean average precision* (mAP) refers to averaging the AP over all classes. When one refers to mAP@0.5, one has fixed an IoU-threshold of 0.5, meaning that one only considers predictions where the IoU metric has value greater than 0.5. The mAP value is the main performance metric used in training object detection models.

The process of training the model is the standard machine learning flow. A large portion of the data set, e.g. 80% is taken as training samples, while the remaining 20% is taken as test samples. We replicate this process in five separate iterations, meaning that we do random sampling into 80-20 split 5 times. Thus we conduct training and testing in 5 different iterations. This is to minimize any bias which would result from having an uneven data split by chance.

### Evaluation of model performance

Our main objective in this work is to train an object detection model. In addition to the standard evaluation metrics such as IoU and mAP, we do the following manual assessment of the predictions on the test set:

▪ Number of detections by human (this is considered ground truth)
▪ Number of correct detections by model (true positive)
▪ Number of incorrect detections by model (false positive)
▪ Number of missing detections by model (false negative)

We also remark that for a dermatologist, the value of the object detection model as a decision support tool does not depend strictly on the model’s ability to give optimal bounding boxes with minimal pixel areas around each acne lesion. For the dermatologist, a fairly accurate bounding box around the acne lesions will suffice. This practical aspect is important to have in mind when evaluating such models. Of course, the IoU and mAP metrics are important in model training and validation, but in practical visual validation the minimization of the area of the bounding box is not essential to model performance and quality.

Hence we work with the following manual evaluation criteria:

| Metric | Description |
| --- | --- |
| Number of detections by model | Number of acne lesions detected by the trained model |
| Number of detections by human | Number of acne lesions detected by the human dermatologist |
| Correct detections (true positive) by model | Number of acne lesions correctly detected by model, as judged by dermatologist |
| Wrong detections (false positive) by model | Number of acne lesions incorrectly detected by model, as judged by dermatologist |
| Missed detections (false negative) by model | Number of acne lesions not detected by model, as judged by dermatologist |

## Results

### Single class detection

The single class (“acne”) detection model achieves a maximum mAP@0.5 value 37.97 after only 5 epochs. We conduct a 5-fold cross validation training with an image size of 640×640 pixels, and a training-test split of 80-20. The training is run for 100 epochs with a batch size of 16 and the SGD optimizer. Each epoch took approximately 2 minutes on an Nvidia Tesla V100 GPU with 16GB VRAM.

| Best mAP@0.5 | Best epoch |
| --- | --- |
| 37.97 | 5 |

| Cross validation fold | Best mAP@0.5 | Best epoch |
| --- | --- | --- |
| 1 | 36.81 | 8 |
| 2 | **37.97** | 5 |
| 3 | 35.22 | 4 |
| 4 | 35.41 | 22 |
| 5 | 36.53 | 20 |

The following evaluation metrics on our clinical data set are achieved using the YOLOv5 model combined with visual inspection of the dermatologist as described above.

| Measure | Value | Explanation |
| --- | --- | --- |
| True positive | 93.59 % | Correct detections |
| False positive | 3.44 % | False detections |
| False negative | 5.20 % | Missed detections |
| Precision | 96.45 % | Fraction of correct detections among all detections: |
| | | $\frac{\text{true positive}}{\text{true positive} + \text{false positive}}$ |
| Recall | 94.73 % | Fraction of correct detections retrieved:<br>$\frac{\text{true positive}}{\text{true positive} + \text{false negative}}$ |
| F1 | 95.58 % | Harmonic mean of precision and recall:<br>$\frac{2 \cdot \text{precision} \cdot \text{recall}}{\text{precision} + \text{recall}}$ |

Note that we do not measure *true negative* (correctly detect non-acne) because healthy skin is not explicitly detected by the model and hence is not relevant.

### Multi class detection

The multi class detection model, where the classes are severity levels 1-4, achieves a maximum mAP@0.5 value 26.50 after 20 epochs. Also, here we use an image size of 640×640 pixels, and a training-test split of 80-20. The training is again run for 100 epochs total with a batch size of 16 and the SGD optimizer. Each epoch took approximately 2 minutes on an Nvidia Tesla V100 GPU with 16GB VRAM.

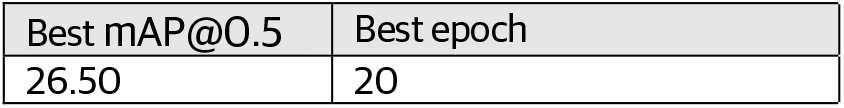

## Discussion

### Model strengths

The YOLOv5 object detection model trained on the ACNE04 has a number of strengths. It is able to be well trained on both the single class task (acne) and the multi class task (0, 1, 2, 3) where the classes are the varying severity levels. As such, the model is able to learn the main different types of acne: papules, pustules, nodules and cysts.

Moreover, the model also seems to correctly learn excoriations and post inflammatory hyperpigmentation and scarring. The model detects acne on all body parts, not only the face. The model is also robust for different skin tones. Blurry images do not disturb the model, and the model accepts varying zoom levels and lighting conditions also.

### Model weaknesses

The trained model does sometimes tend to confuse certain body regions for acne lesions. These body regions are typically the creese of the lips, nostrils, nipples and sometimes hair follicles. The reason is evidently that these body parts do have similar shape and color to inflammatory acne lesions, i.e. having a red or pink color. Another source of confusion are hyperpigmented lesions, as acne too is often accompanied with post-inflammatory hyperpigmentation. We hypothesize that these confusions can be alleviated by introducing more photos of such artefacts without acne, so that the model can learn by seeing more complicated true negative samples.

## Conclusions

Our object detection model is trained on a small scale publicly available data set and is proving to be useful in our clinical practice. We have shown that one can achieve fairly accurate models by simple means.

The mAP metrics have much room for improvement, but the model is still useful at this performance level. We also note that the training of standard architecture object detection models has given significantly better accuracy than the more intricate custom approaches in the existing research literature concerning acne detection in photographs

## Data Availability

All data in the ACNE04 dataset is publicly available online.

https://github.com/xpwu95/LDL

## Notes

### Competing Interest Statement

The authors are employed by the private digital health care company Askin, providing teledermatological services in Norway.

### Funding Statement

This study did not receive any funding.

### Author Declarations

The ACNE04 dataset is publicly available on the internet site https://github.com/xpwu95/LDL

